# Using theater-based research to study views on contraception

**DOI:** 10.1101/2025.04.10.25325356

**Authors:** Wieke Y. Beumer, Merel Sprenger, Ebru Eksi, Chris Bartels, Samira van Bohemen, Jenneke van Ditzhuijzen, Linda P. Dekker, Ingvil de Haan, Hanan El Marroun, Pauline W. Jansen, Hilmar H. Bijma, Clair A. Enthoven

## Abstract

**Objectives:** Historically, contraceptive research is impeded by barriers to openly discuss sexual experiences. Theater-based research is a promising approach to foster deeper engagement and a richer exploration of lived experiences to gain insights in underlying implicit factors that shape contraceptive use. This study uses theater-based research to explore adults’ views, responsibility, satisfaction, and knowledge about contraception.

**Study Design:** The study was conducted at a three-day music festival in the Netherlands with 1,024 participants (32.9% men, mean age 28.1 years). Theater was integrated by transforming the study setting into a dance club, where the research team performed characters. Participants completed an individual survey prior to the theater experience, a team-based contraceptive knowledge quiz, and a writings on the wall activity. Survey and quiz data were analyzed using linear regression, thematic analysis was applied for evaluating writings on the wall.

**Results:** On a scale from 1 to 10, the survey revealed general contraceptive satisfaction (women: 7.0, men: 7.7) and responsibility (women: 8.7, men: 7.7). Participant teams had a mean of 73.7% correct quiz answers. We found six themes that emerged in contraceptive views: side effects, fallibility, shared responsibility, information needs, dismissive counseling, and resistance to hormones.

**Conclusion:** This study revealed complementary findings between traditional survey findings, indicating overall satisfaction with contraception, and theater-based writings on the wall, eliciting more candid and critical reflections with a notably negative tone. Theater-based data collection may capture lived experiences more vividly and lower barriers for research participation, suggesting its value for studying sexual and reproductive health.

**Implications:** Theater offers a promising complement to conventional contraception research. By fostering comfort, humor and participation, it reveals more candid perspectives and may therefore reduce social desirability bias. Future research should further examine its added value in recruiting populations not reached by traditional methods.

## 1 Introduction

Contraceptive methods play a central role in empowering people to make autonomous reproductive choices (Alspaugh et al., 2020). In settings where contraceptive care is widely available and generally accessible, discussions have focused on the reasons for a recent decline in (hormonal) contraceptive use (de Graaf et al., 2024; Le Guen et al., 2021; Schneider-Kamp & Takhar, 2023). A substantial body of qualitative research, primarily using semi-structured interviews and focus groups, shows that contraceptive use is not only shaped by the desire to avoid pregnancy, but also by negative views, satisfaction, and (sexual) partner dynamics (Alspaugh et al., 2020; Ayoola et al., 2007; Baxter et al., 2011; D’Souza et al., 2022; Hoga et al., 2014; Pratt et al., 2014; Williamson et al., 2009). These implicit factors may not be fully understood using traditional research methods, such as surveys, interviews and focus groups, as social desirability may limit gauging the lived experience of people, particularly when exploring intimate experiences and personal perspectives, such as contraception (Tourangeau & Yan, 2007). This complexity underscores the need for novel methodological approaches.

An approach to elucidate and evaluate implicit aspects of people’s lived experiences is arts-based research. Arts-based research draws on diverse ways of knowing and experiencing the world (Knowles & Cole, 2008) and serves as an umbrella term encompassing a range of methodologies guided by constructivist, experiential, and empirical principles. Examples of arts-based methods include poetry, photography, painting, and theater: art forms that create interactions among participants extending beyond structured conversations, fostering deeper emotional and cognitive engagement and encouraging self-reflection (Myllyoja, 2024).

Theater-based research is an arts-based method in which theatrical elements are used during data collection (such as improvisation, theatrical costumes and role play), to lower participation barriers and create an open, playful environment that enables participants to articulate experiences and perspectives that might otherwise remain unspoken (Bleuer et al., 2018; Collins & Stockton, 2022; Myllyoja, 2024; Rossiter et al., 2008). Theater-based research may be particularly useful in research on contraception because it has the potential to engage a broader and typically less included sample (Knowles & Cole, 2008), addressing the underrepresentation of men commonly observed in contraception studies (Alspaugh et al., 2020). Furthermore, theater promotes knowledge exchange, allowing participants to gain new insights through active involvement in the research process (Bleuer et al., 2018; Myllyoja, 2024). As a result, theater has thus far been used mainly as a medium for disseminating knowledge (Bleuer et al., 2018; Fleckman et al., 2023; Marzi et al., 2025; Rossiter et al., 2008), while its application as a method for data collection remains largely unexplored. Therefore, in this study we aim to explore contraceptive views, responsibility, satisfaction and knowledge among adult women and men using theater-based research.

## 2 Methods

### 2.1 Setting and participants

Participants aged 18 or older were recruited at Lowlands, a large yearly recurring music festival attracting around 65,000 visitors in the Netherlands during three days in August 2023. Besides music, the festival offers a wide variety of entertainment including theater, movies, comedy, literature, visual arts and science. Lowlands Science is an initiative to foster science engagement by welcoming ten different research projects at Lowlands each year (Docter-Loeb, 2024). The participants provided digital informed consent, and individuals who were noticeably intoxicated could not participate. The protocol received approval from the Ethics Review Committee of Erasmus School of Social and Behavioral Sciences (ETH2223-0634). Our research project entitled: “Perception of Contraception”.

### 2.2 Design

This study included an individual online survey, a team-based contraceptive knowledge quiz and a writings on the wall activity. The online survey and quiz were co-created with stakeholders, including a gynecologist and two program managers working at a societal organization to promote sexual and reproductive health. The survey was conducted prior to the interactive theater experience, the quiz during the theater experience and the writings on the wall at the end of this experience (See next section ‘Interactive theater’).

The survey was conducted on participants’ smartphones and included questions about age, sex, gender, educational attainment, sexual orientation, presence of a steady sexual partner (hereafter referred to as ‘relationship’), and number of sexual partners in the past year. Participants then indicated whether they did anything to prevent pregnancy (hereafter referred to as ‘contraceptive use’) and which methods they used. Finally, they answered questions about satisfaction with their contraception and their sense of responsibility on a scale from 0 (not at all) to 10 (very much). The survey questions and answer options are described in Supplementary Table 1.

The contraceptive knowledge quiz was conducted among small teams (two to six participants per team). The survey included eight questions with four answer options, based on existing questions (Haynes et al., 2017; Sense). Topics included effectiveness and working mechanisms of contraceptive methods, protection against sexually transmitted infections (STIs), sperm viability, and the fertile period. A full description of the quiz is shown in Supplementary Table 2X.

The writings on the wall consisted of wall panels featuring the invitation, “What is your opinion about…” and contained large drawings of contraceptive methods, allowing participants to write about these specific methods. The depicted contraceptive methods included: the contraceptive pill, injection, ring, or patch, the condom, internal condom, diaphragm, (hormonal) intrauterine device (IUD), and sterilization. Empty spaces around these drawings provided an opportunity to describe views toward contraception in general.

### 2.3 Interactive theater

The theater design was developed during multiple one-on-one brainstorm sessions between one of the researchers (CAE) and actors with extensive experience with development and performance of interactive theater acts at festivals. Theater-based aspects of the study included the use of costumes, role play, improvisation, attributes for participants to interact with, and a research area designed to create a strong visual impact.

We created a dance club named ‘Club Womb’ as our project setting, visually presented in Figure 1. At the entrance, the ‘bouncer’ explained the purpose of the study, organized digital consent and directed participants towards a brief online survey (Figure 1.1). Next, the ‘host’ welcomed the participants, explained that they could “protect themselves from pregnancy” during their stay in ‘Club Womb’ and invited them to choose one of ten enlarged contraceptive methods (e.g.., raincoat as condom, hoop as contraceptive ring; Figure 1.2). Equipped with their method of choice, participants entered the club by crawling through a large vulva after touching the ‘clitoris’ (a plasma ball light; Figure 1.3). Inside, the ‘sexual health doctor’ welcomed participants in the “most visited womb of the festival”, conducted a contraceptive knowledge quiz and explained the correct answers (Figure 1.4). Finally, participants moved into the silent disco where the ‘DJ/MC’ (Figure 1.5) invited them to write their opinions and experiences with contraceptive methods on the wall (Figure 1.6). The club ‘mascot’ (actor in a penis suit) recruited participants and danced with them (Figure 1.1). Throughout the experience, all actors helped participants feel at ease by using empathetic communication and humor, and by adapting their approach to suit individual comfort levels.

**Figure 1.**
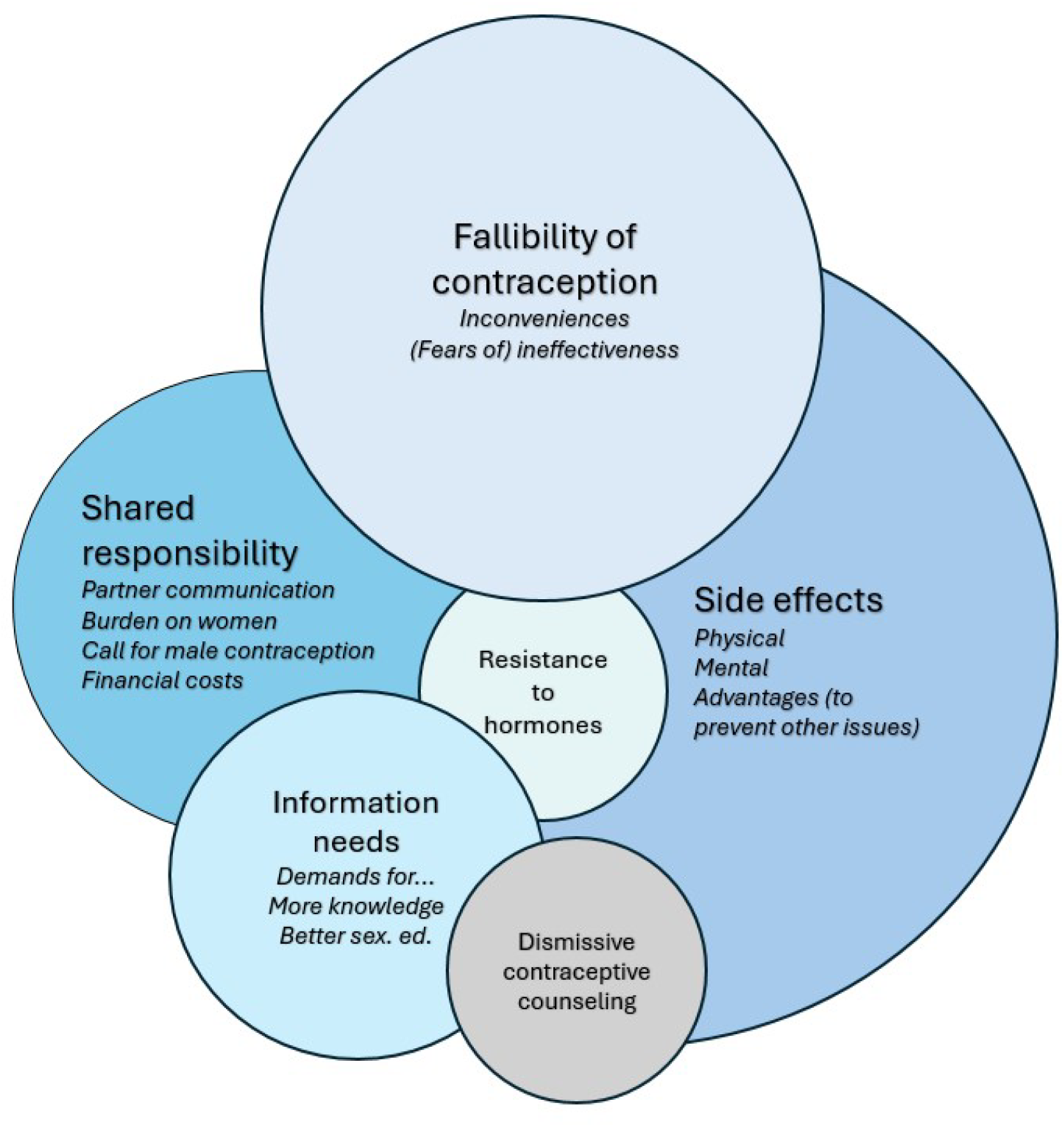
The stages of the study at Lowlands festival, The Netherlands, 2023.

Throughout data collection, we continuously reflected on how participants engaged with the theatrical aspects of the study and the specific characters performed by the actors. Observations focused on participants’ movement through the three stages and on identifying possible improvements to the design and interactions. Team reflection sessions were held four times—once midway through the first day and at the end of each of the three days. Reflections on the benefits and drawbacks of the use of theater-based research from three researchers (WB, MS, CAE) are described in the discussion section.

### 2.4 Analyses

Regarding the quantitative analyses, for the online survey, means and standard deviations were computed for contraceptive satisfaction and responsibility separately for women and men. For the contraceptive knowledge quiz, the mean and standard deviation of the percentage of correct answers per team were calculated separately for female, male, and mixed teams. We additionally conducted multivariate linear regression analyses with sex as explanatory variable and contraceptive satisfaction, responsibility, and knowledge as outcome variables, adjusted for age, educational attainment, and relationship. We used IBM SPSS Statistics, Version 28.0.1.0.

Regarding the qualitative analyses, for the writings on the wall, we photographed the panels and transcribed them verbatim into MS Excel for inductive data processing and analysis. Besides verbatim contributions, participants interacted with other quotes through arrows or additional punctuation marks. We used thematic analysis in our exploratory approach of identifying, analyzing, and reporting of themes within the data (Braun & Clarke, 2006). Data extraction, open coding, axial coding, and constructing the overarching themes was performed by CB, in consultation with CAE. Subsequent analyses were performed by WB. Since this is an iterative process, the researchers went back and forth between axial coding and constructing the themes. Decisions were made based on consensus within the team (CB, CAE and WB).

## 3 Results

### 3.2 Sample characteristics

A total of 1,088 people started the study. We excluded 64 participants due to incomplete data, leaving 1,024 participants. 744 Participants, divided over 292 teams, also participated in the contraceptive knowledge quiz. Participants had a mean age of 28.1 years (range 18 to 62) and 33.6% identified as male. The majority of the participants had a high educational attainment and were in a relationship (Tables 1 and Supplementary Table 2). Most participants (91.3%) reported using contraception. Of those who did not use contraception, most (73.0%) were either not engaging in penis-in-vagina sex, were pregnant, or were trying to conceive. When contraception was used, the hormonal IUD, condoms, and the contraceptive pill were most commonly reported (Supplementary Table 3).

**Table 1.** Characteristics (n(%)) of the study sample (n=1,024) at Lowlands festival, The Netherlands, 2023.

|  | Total sample<br>n=1,024 | Women<br>n=683 | Men<br>n=341 |
| --- | --- | --- | --- |
| **Age** |  |  |  |
| 18-24 years | 288 (22.3) | 161 (23.6) | 67 (19.6) |
| 25-29 years | 497 (48.5) | 354 (51.8) | 143 (41.9) |
| ≥30 years | 299 (29.2) | 168 (24.6) | 131 (38.4) |
| **Gender** |  |  |  |
| Female | 677 (66.1) | 677 (99.1) | 0 |
| Male | 344 (33.6) | 3 (0.4) | 341 (100) |
| Other | 3 (0.3) | 3 (0.4) | 0 |
| **Educational attainment<sup>1</sup>** |  |  |  |
| Low | 5 (0.5) | 1 (0.1) | 4 (1.2) |
| Medium | 158 (15.4) | 95 (13.9) | 63 (18.5) |
| High | 857 (83.7) | 585 (85.7) | 272 (79.8) |
| Prefer not to say | 4 (0.4) | 2 (0.3) | 2 (0.6) |
| **Usually has sex<sup>2</sup> with** |  |  |  |
| Someone from the opposite sex | 910 (88.9) | 600 (87.8) | 310 (90.9) |
| Someone from the same sex | 82 (8.0) | 56 (8.2) | 26 (7.6) |
| Both | 25 (2.4) | 24 (3.5) | 1 (0.3) |
| Never had sex<sup>2</sup> | 6 (0.6) | 3 (0.4) | 3 (0.9) |
| Not listed | 1 (0.1) | 0 | 1 (0.3) |
| **Relationship** |  |  |  |
| Yes | 768 (75) | 488 (71.4) | 280 (82.1) |
| No/unsure/prefer not to say | 256 (25.0) | 195 (28.6) | 61 (17.9) |
| **Number of sexual<sup>2</sup> partners in past year** |  |  |  |
| **≤ 1** | 678 (66.2) | 435 (63.7) | 244 (71.6) |
| **2-10** | 325 (31.8) | 236 (34.6) | 89 (26.1) |
| **>10** | 18 (1.8) | 10 (1.5) | 8 (2.3) |
| **Using contraception** |  |  |  |
| **No** | 89 (8.7) | 50 (7.3) | 39 (11.4) |
| **Yes** | 935 (91.3) | 633 (92.7) | 302 (88.6) |
| **If yes, which method<sup>3</sup>** |  |  |  |
| **External condoms** | 350 (37.4) | 222 (35.1) | 128 (42.4) |
| **Hormonal IUD** | 311 (33.3) | 223 (35.2) | 88 (29.1) |
| **Contraceptive pill** | 253 (27.1) | 175 (27.7) | 78 (25.8) |
| **I make sure my partner uses contraception** | 28 (3.0) | - | 28 (9.3) |
| **Cycle tracking** | 41 (4.4) | 32 (5.1) | 9 (3.0) |
| **Copper IUD** | 73 (7.8) | 49 (7.7) | 24 (7.9) |
| **Withdrawal** | 48 (5.1) | 30 (4.7) | 18 (6.0) |
| **Morning after pill** | 27 (2.9) | 20 (3.2) | 7 (2.3) |
| **Contraceptive implant** | 15 (1.6) | 9 (1.4) | 6 (2.0) |
| **Sterilization** | 16 (1.7) | 8 (1.3) | 8 (2.6) |
| **Contraceptive/vaginal ring** | 12 (1.3) | 7 (1.1) | 5 (1.7) |
| **Internal condoms** | 7 (0.7) | 5 (0.8) | 2 (0.7) |
| **Contraceptive injection** | 4 (0.4) | 2 (0.3) | 2 (0.7) |
| **Contraceptive patch** | 3 (0.3) | 2 (0.3) | 1 (0.3) |
| **Diaphragm** | 1 (0.1) | 0 (0.0) | 1 (0.3) |

<sup>1</sup> Low: primary education or pre-vocational secondary education; Medium: lower/upper general secondary education, or secondary vocational education; High: higher professional education or university.
<sup>2</sup> Sex is broadly defined, such as manual sex, oral sex, penis-in-vagina sex, or anal sex.
<sup>3</sup> Multiple answers were allowed. Participants were asked about their and/or their partner's contraceptive methods, which is why men also reported contraceptives such as the contraceptive pill or IUDs. Given that some participants in the study were couples, there might be overlapping data in contraception methods. Cycle tracking includes periodic abstinence, calendar method, no penis-in-vagina sex mid-cycle, or using a fertility app.

### 3.2 Contraceptive satisfaction, responsibility, and knowledge

The mean satisfaction score of those using contraception (n=935) was 7.2 out of 10. Both women and men rated their contraception as satisfactory, with women being significantly less satisfied than men with their current method (Table 2). The mean sense of responsibility (n=1,024) was 8.4 out of 10. Women reported a significantly higher sense of responsibility towards contraceptive use compared to men (Table 2). The mean percentage of correct answers in the contraceptive knowledge quiz was 73.7%. Of all teams, 94.5% had 50% or more correct answers (Supplementary Table 3). The all-male teams had significantly lower knowledge (61.1%) than all-female (77.5%) or mixed (74.6%) teams (Table 2).

**Table 2.**
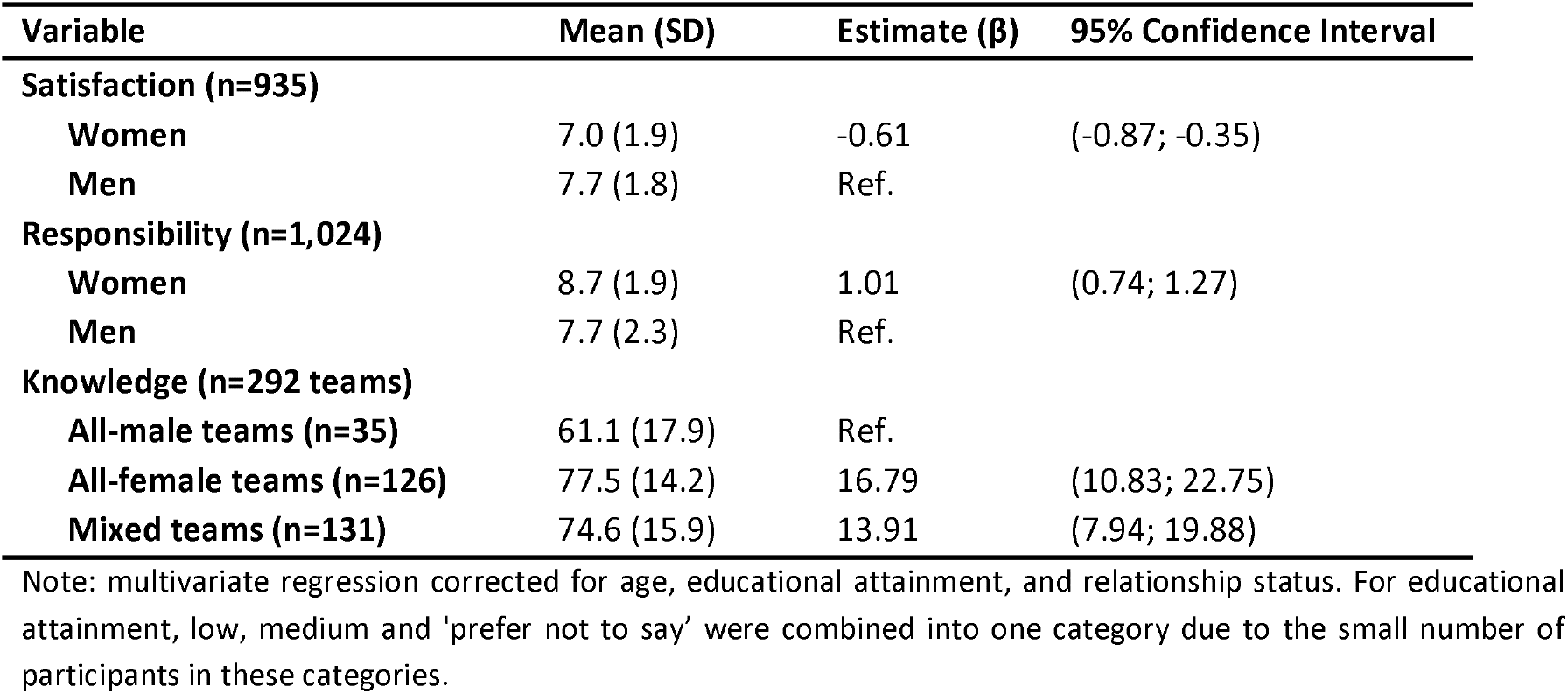
Mean scores of satisfaction, responsibility and knowledge for women and men separately at Lowlands festival, The Netherlands, 2023.

### 3.3 Contraceptive views

We identified five themes (Figure 2). We defined the themes broadly, reflecting the exploratory nature of the study. The thematization is not intended to be rigid or mutually exclusive but aims to provide a framework for understanding the range of diverse views. Illustrative quotes per theme are displayed in Table 3.

**Figure 2.** Schematic overview of overlapping themes in the writings on the wall at Lowlands festival, The Netherlands, 2023. Larger circles represent themes that were mentioned more frequently and indicate greater overlap. Subthemes are in italics.

**Table 3.**
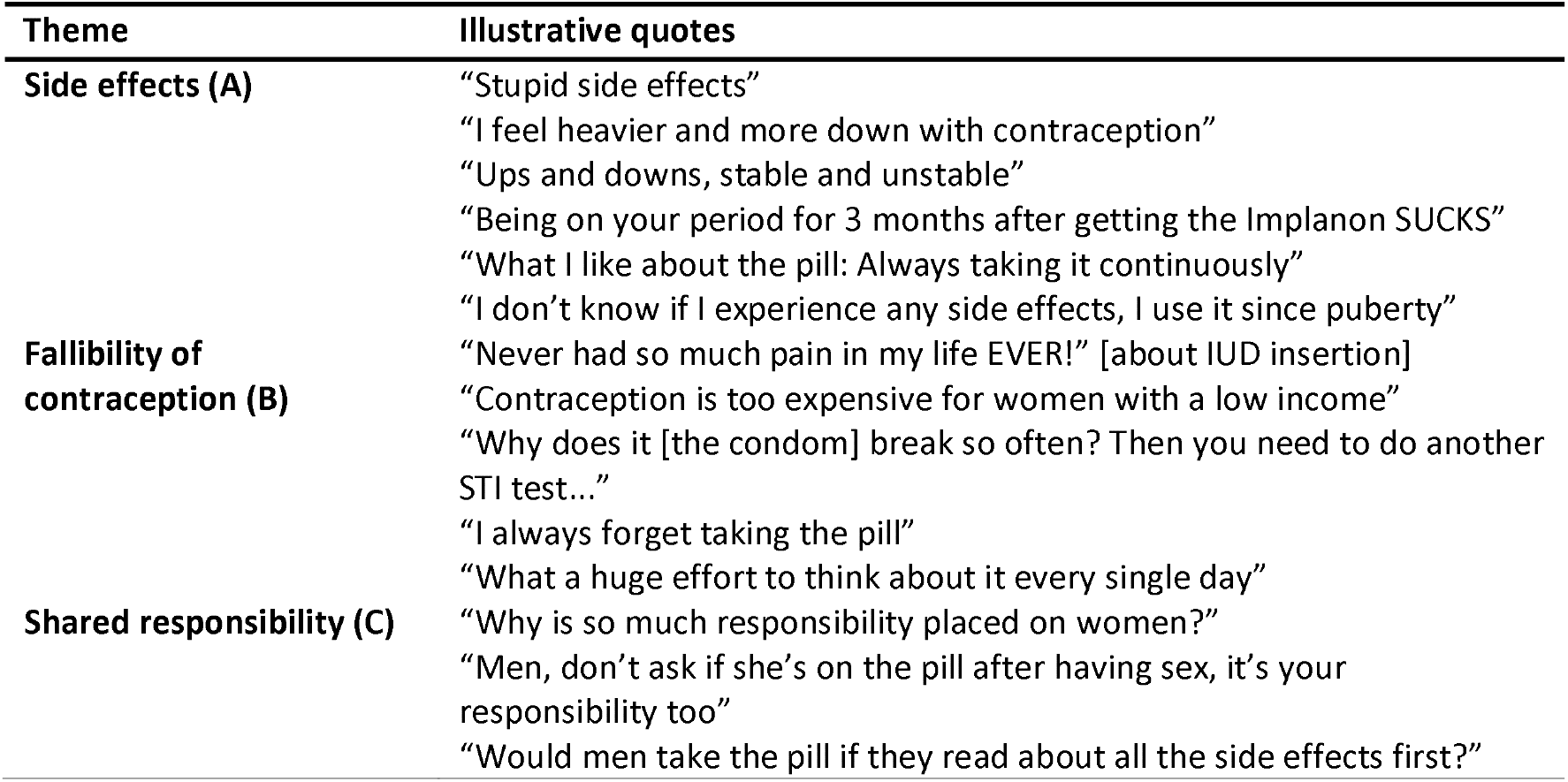
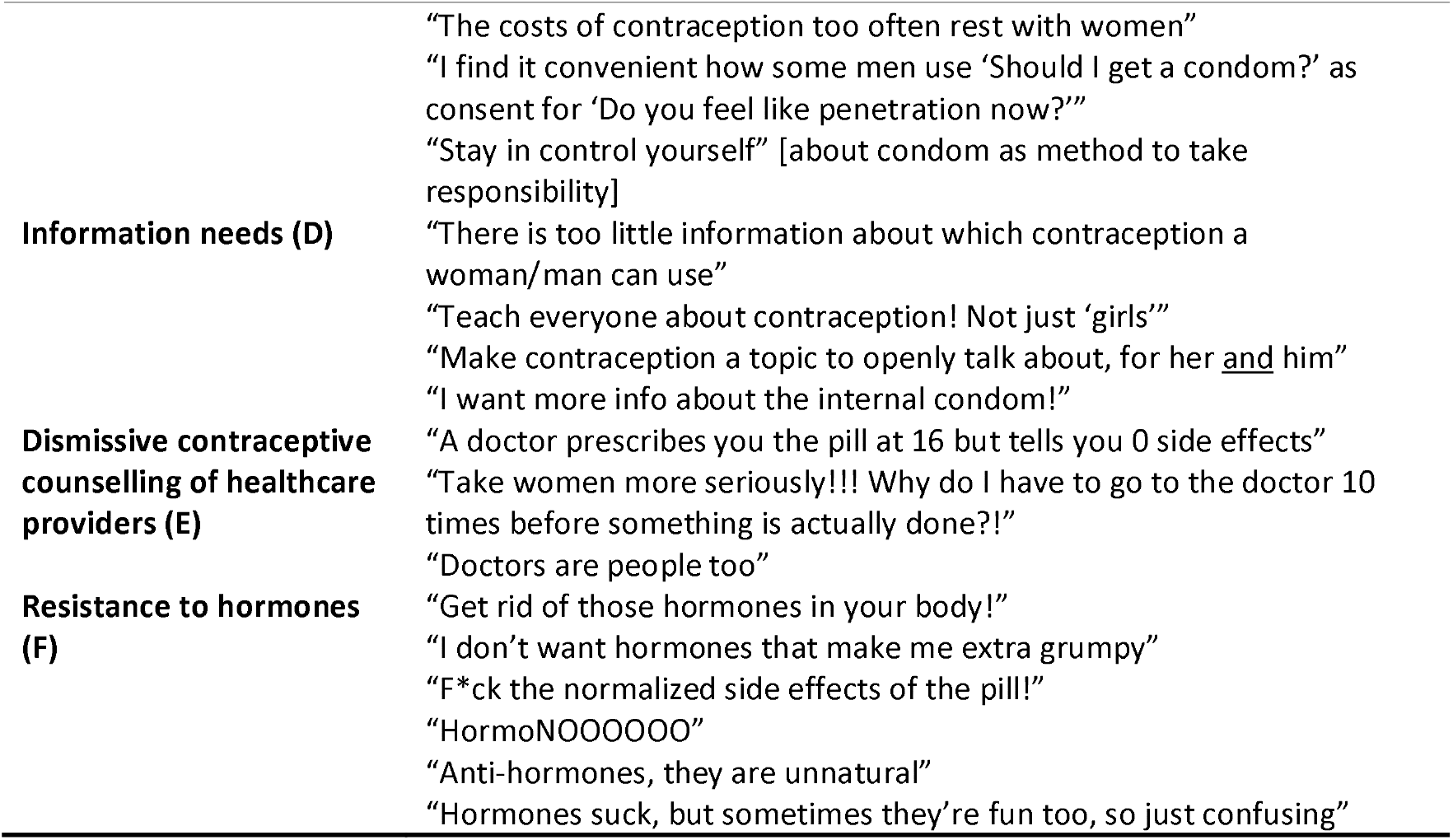
Illustrative quotes for each theme from the writings on the wall activity at Lowlands festival, The Netherlands, 2023.

Many participants expressed concerns about physical and mental **side effects** (Theme A) of contraception, often in a negative tone. These included general worries or personal experiences such as mood swings and heavier bleeding. At the same time, some noted perceived benefits, like menstrual regulation and reduced acne.

Various practical challenges related to the **fallibility of contraception** (Theme B) were also mentioned. Condoms were often described as unpleasant or unromantic interruptions during sex, and some participants reported issues with effectiveness, such as breakage or leakage. Other inconveniences included the painful insertion of an IUD, high costs of contraception, and forgetfulness— like missing a pill.

Further, there was a strong call for more **shared responsibility** (Theme C) in contraceptive use. Participants felt that the burden of side effects and costs disproportionately affects women. They emphasized the importance of open communication and shared decision-making. Some advocated for male contraceptive options, though others were skeptical about men’s willingness to use them. Positive examples of shared responsibility were also mentioned, such as using condom negotiation as a way to signal consent.

Participants also expressed a clear **need for information** (Theme D). Questions were raised about lesser-known methods like the internal condom and diaphragm, and many felt underinformed about side effects. There was also a call for more research into non-hormonal alternatives and for improved comprehensive sexuality education.

Participants also described **dismissive attitudes from healthcare providers during contraceptive counseling** (Theme E). Others shared frustrations about not being taken seriously when seeking advice, such as a participant whose concerns about an implant were dismissed with a referral to a psychologist.

Finally, participants voiced **resistance to hormonal methods** (Theme F), citing experienced side effects or perceived long-term risks, such as potential impacts on fertility. These writings were often related to other themes. For instance, some participants emphasized the importance of having their concerns about hormonal contraception taken seriously by HCPs.

## 4 Discussion

In this study, we applied theater-based data collection to study contraception. Our different data collection methods complemented each other: while the quantitative methods showed sufficient satisfaction, responsibility and knowledge, the writings on the wall revealed that participants also perceived contraception as suboptimal due to (hormonal) side effects, the unequal burden on women, and a lack of recognition in contraceptive concerns from healthcare providers.

Prior literature has suggested that drama methods may provide deeper insights into lived experiences and embodied understanding (Myllyoja, 2024). While participants initially rated their contraception as satisfactory in the survey, conducted outside the theater setting, more nuanced (and primarily negative) views appeared on the writings on the wall towards the end of the study, after the interaction with actors. The themes that emerged in the writings on the wall align with findings from previous studies that used more traditional research methods, such as interviews (Alspaugh et al., 2020; Ayoola et al., 2007; Baxter et al., 2011; Brown, 2015; D’Souza et al., 2022; Gollub et al., 2022; Hoga et al., 2014; Kimport, 2018; Manzer et al., 2023; Pratt et al., 2014; Stewart et al., 2017; Wagner et al., 2024; Williamson et al., 2009). However, compared to traditional data, participants’ contributions to our writings on the wall were noticeably more candid and nuanced – as if they were talking among friends – thereby unveiling their lived experiences more vividly. For example, the language used in the writings on the wall was informal and emotionally charged (for instance: “HormoNOOOO”), which may not be as effectively captured in formal interview settings.

To the authors’ knowledge, this is the first study to use a theater-based approach as a way to evaluate views and lived experiences of contraception, yielding both quantitative and qualitative insights in a group of over a thousand participants. Given its novelty, we will describe some reflections on the use of theater as a data collection method to facilitate replication and support other researchers who consider using this method. Firstly, we noticed that a theater-based approach offers unique ways to increase recruitment and engagement. By making it fun, we lowered barriers for participation. This resulted in some participants who were initially hesitant during recruitment– particularly men, who sometimes felt they did not belong in research on contraception – to also take part. Ultimately, we attracted a large number of participants, of whom one third were men. Through improvisation within their character, the research team was able to tailor interactions to each individual, for example by tapping into their curiosity about the theater setting of Club Womb, exploring why their participation was relevant. Watching other festival visitors enter the experience, selecting enlarged contraceptive methods and crawling through the door-sized vulva, in ponchos as condoms, increased the curiosity about what was awaiting inside Club Womb, our theater research area. As the entrance also served as the exit, festival visitors could see returning participants smiling and excitedly chatting about the study. Meanwhile, the ‘mascot’, moving back and forth between the silent disco and the festival area, drew festival visitors’ attention and further increased participation. The setting was therefore very attractive for participants to take part in the study.

Secondly, we notice an increased candor of participants to share their views and experiences (Bleuer et al., 2018; Collins & Stockton, 2022; Myllyoja, 2024; Rossiter et al., 2008). The theatrical playfulness visibly helped participants feel at ease, as we noticed them chatting to strangers already at the start, when the host was welcoming them into the experience. As the study progressed, participants became increasingly open, for example during the knowledge quiz, they laughed at the sexual health doctor’s jokes and started sharing personal experiences. When they then entered the club area, participants danced together while reflecting on their contraceptive views and experiences. They interacted with each other in their writings and drawings, and by talking to each other. Humorous one-liners of the ‘DJ/MC’ amplified the sense that there were no limits on what could be shared. The text on the writings on the wall showed that participants used their own language, as if among friends, which they might less likely use in an in-depth interview. Therefore, theater-based data collection may help reduce social desirability bias.

Our study shows the potential of theater-based research in evaluating contraceptive views, evoking more open and authentic responses than is possible in traditional research. Nevertheless, this study also has some limitations. As a result of our study setting, our sample consisted of relatively highly educated festival participants and was therefore not representative of the Dutch population. Additionally, results from the contraceptive knowledge quiz reflect shared rather than individual knowledge and may therefore overestimate actual knowledge levels. Finally, as we did not collect data on the participants’ experiences of the theater-based research methods – wanting to first pilot this method rather than setting up a formal evaluation in addition to what we were already asking of participants – we were unable to provide an evaluation beyond our own perspectives as researchers and actors in the project. More research is needed to further investigate the potential contribution of theater-based data collection to research, especially regarding 1) the inclusion of populations that are not reached by traditional research methods, and 2) the type of research questions for which this type of data collection is most suitable.

## 5 Conclusion

This study applied theater-based data-collection to explore contraceptive views among adult women and men. Regarding contraceptive satisfaction, the survey, conducted outside the theater-setting, and the writings on the wall, towards the end of the theater-setting, complemented each other. While participants appeared relatively satisfied with their contraception in the survey, the writings on the wall evolved into a collection of candid, critical expressions, highlighting negative experiences with various aspects of contraception, suggesting clear points for improvement. We believe theater may have further encouraged participants to express themselves in their own words, as shown on the writings on the wall. Theater-based data collection may therefore complement traditional research, fostering more inclusive, engaging, and dynamic investigations of sexual and reproductive health, while capturing people’s lived experiences.

## Data Availability

All data produced in the present study are available upon reasonable request to the authors.

## Acknowledgements

We gratefully acknowledge Lowlands Science 2023 for the organization. In addition, we would like to acknowledge all participants who took the time to participate in research while attending a music festival. Furthermore, we would like to thank Ineke van der Vlugt, program manager Rutgers, Expert Centre on Sexuality, for providing feedback on the questionnaire and knowledge quiz. Finally, we want to acknowledge the research team for their creativity in designing this project and for the energy they brought in the execution of the project during the festival weekend: Bart, Eveline, Jan, Nena, Rosa, Saskia, Teun, and Vincent.

## Data availability statement

Anonymized survey, quiz data and syntaxes are openly available in the EUR Data Repository at (https://figshare.com/s/cc145e475ad1ba294d14).

## Ethical statement

All participants provided digital informed consent, and the research protocol received approval from the Ethics Review Committee of Erasmus School of Social and Behavioral Sciences (ETH2223-0634).

