## Appendix A for "Using theater-based research to study views on contraception"

### Appendix A of manuscript: Contraceptive views, satisfaction, responsibility, and knowledge: A theater-based study at the Dutch 'Lowlands' festival

**Supplementary Table 1.** *Description of survey measures at Lowlands festival, The Netherlands, 2023.*

| **Measure** | **Survey item and answer options** | **Description** |
|---|---|---|
| **Age** | How old are you?<br>*[Numerical]* | Age ranged from 18 to 62, with a mean age of 28.1 and a positively skewed distribution. Therefore, the following categories were created: 18-24 years, 25-29 years, and ≥30 years.<br><br>For the teams participating in the knowledge quiz, mean age for each team was computed and categorized into the same categories as described above. |
| **Sex** | What is your biological sex?<br>*Man*<br>*Woman*<br>*Different*<br>*Prefer not to say* | For sex assigned at birth, in the article referred to as sex, the categories men and women were used, since none of our participants filled out 'different' or 'prefer not to say.'<br><br>For the teams participating in the knowledge quiz, we categorized the teams in the following three categories: all-male, all-female, and mixed. |
| **Gender** | How do you see yourself?<br>*Man*<br>*Woman*<br>*Non-binary*<br>*Other*<br>*Prefer not to say* | Gender was only used for descriptive statistics, and consisted of the following three categories: man, woman, and other (consisting of other and non-binary). |

| | | |
|---|---|---|
| **Educational attainment** | What is the highest level of education you have completed?<br>*Primary education or part of it*<br>*Pre-vocational secondary education (VMBO-vocational track, LBO, or practical education)*<br>*Lower general secondary education (MAVO, VMBO-theoretical/mixed track, (M)ULO, middle school, first three years of HAVO or VWO)*<br>*Secondary vocational education (MBO)*<br>*Upper general secondary education (HAVO, Atheneum, VWO, Gymnasium)*<br>*Higher professional education (HBO)*<br>*University education (WO)*<br>*Other, namely: ...*<br>*Prefer not to say* | Educational attainment was categorized into:<br>Low: primary education or pre-vocational secondary education;<br>Medium: lower/upper general secondary education, or secondary vocational education; and<br>High: higher professional education or university education<br>Prefer not to say<br><br>Due to the small number of respondents in the 'low' and 'prefer not to say' categories, these groups were combined with the 'medium' category.<br><br>For the teams participating in the knowledge quiz, educational attainment was divided into three categories: only low or only medium, only high, and mixed. |
| **Usually has sex with** | If you have sex*, is it usually with...<br>*Someone of the same biological sex*<br>*Someone of a different biological sex*<br>*Both*<br>*I have never had sex*<br>*Other, namely: ...*<br>*Sex can mean many things, such as oral sex, fingering, hand jobs, penis-in-vagina sex, or anal sex. | This variable was used for descriptive statistics and was categorized in the following groups based on biological sex: someone from the opposite sex, someone from the same sex, both, never had sex, and not listed. |
| **Relationship** | Do you currently have a steady sexual partner?<br>*Yes*<br>*No*<br>*I don't know*<br>*Prefer not to say* | Presence of a steady sexual partner, referred to as 'relationship' was divided into two categories: yes and no/unsure/prefer not to say.<br><br>For the teams participating in the knowledge quiz, team relationship status was divided into three categories: single only, relationship only, and mixed. |

| | | |
|---|---|---|
| **Number of sexual partners** | How many sexual partners have you had in the past year? By sexual partner, we mean someone with whom you have had sex*. If you're not sure, please provide an estimate:<br>*[Numerical]*<br>*Sex can mean many things, such as oral sex, fingering, hand jobs, penis-in-vagina sex, or anal sex. | The number of sexual partners was a numerical variable and was divided into three categories: ≤ 1, 2-10, and >10 . |
| **Contraceptive use** | Do you or your sexual partner do anything to prevent pregnancy?<br>*Yes*<br>*No* | The categories for this variable were the same as in the survey: yes and no. |
| **Contraceptive method** (if using contraception, male sex, no steady sexual partner) | What do you or your partner use/do to prevent pregnancy? Multiple answers are possible.<br>*External condoms*<br>*I make sure I know that my partner is using contraception*<br>*I am or my partner is sterilized*<br>*Withdrawal (pulling out)*<br>*I do not have penis-in-vagina sex* | The two questions on contraceptive methods were combined. Since multiple answers were possible, each answer option was a variable coded with either 0, if it was not selected, or 1, if it was selected. The numbers and percentages were calculated for the following methods:<br>External condoms<br>Hormonal IUD<br>Contraceptive pill<br>I make sure my partner uses contraception<br>Cycle tracking (period abstinence or fertility app)<br>Copper IUD<br>Withdrawal<br>Morning-after pill<br>Contraceptive implant<br>Sterilization<br>Contraceptive/vaginal ring<br>Internal condoms<br>Contraceptive injection<br>Contraceptive patch<br>Progestin-only pill (mini-pill)<br>Diaphragm |

| | | |
|---|---|---|
| **Contraceptive method**<br>(if using contraception and previous item not shown) | What do you or your partner use/do to prevent pregnancy? Multiple answers are possible.<br>*The pill*<br>*The mini pill*<br>*External condoms*<br>*Internal condoms*<br>*The contraceptive injection*<br>*The contraceptive patch*<br>*The contraceptive ring (NuvaRing)*<br>*The contraceptive implant (Implanon)*<br>*Copper IUD (non-hormonal IUD)*<br>*Hormonal IUD*<br>*The diaphragm*<br>*Periodic abstinence/calendar method/avoiding penis-in-vagina sex during certain times in the cycle*<br>*I use a fertility app*<br>*Withdrawal (pulling out)*<br>*I do not have penis-in-vagina sex*<br>*I am or my partner is sterilized*<br>*Morning-after pill*<br>*Other, namely: ...*<br>*I don't know*<br>*Prefer not to say* | |
| **Contraceptive satisfaction** | How satisfied are you at the moment with the contraceptive method(s) you are using?<br>*Slider from 0 to 10* | This variable was treated as a continuous variable. |
| **Contraceptive responsibility** | How responsible do you feel to use contraception?<br>*Slider from 0 to 10* | This variable was treated as a continuous variable. |
| **Contraceptive knowledge** | See Supplementary Table 2 for the eight contraceptive knowledge quiz questions. | For contraceptive knowledge, we computed the percentage of correct answers, by dividing the number of correct answers by 8 and multiplying it by 100. This was then treated as a continuous variable. |

**Supplementary Table 2.** *Characteristics of the teams participating in the knowledge quiz (n=292 teams consisting of a total of n=744 individuals) at Lowlands festival, The Netherlands, 2023.*

| **N=292** | **Total sample, n (%)** |
|---|---|
| **Mean age of the teams** | |
| **18-24 years** | 59 (20.2) |
| **25-29 years** | 160 (54.8) |
| **≥30 years** | 73 (25.0) |
| **Sex of the teams** | |
| **All-male** | 35 (12.0) |
| **All-female** | 126 (43.2) |
| **Mixed** | 131 (44.9) |
| **Team educational attainment** | |
| **Only low or only medium** | 18 (6.2) |
| **Only high** | 196 (67.1) |
| **Mixed** | 78 (26.7) |
| **Relationship status of the teams** | |
| **Single only** | 17 (5.8) |
| **In relationship only** | 159 (54.5) |
| **Mixed** | 116 (39.7) |

**Supplementary Table 3.** *Contraceptive knowledge quiz questions and distribution of answers at Lowlands festival, The Netherlands, 2023. Correct answers are in bold.*

| Questions | n (%) |
|---|---|
| 1. Which contraceptive method is the most reliable? | |
| The contraceptive injection | 68 (23.3) |
| **The contraceptive implant** | **154 (52.8)** |
| Oral contraception | 69 (23.6) |
| The contraceptive patch | 1 (0.3) |
| 2. Which of the following contraceptive methods without hormones is the most reliable? | |
| The diaphragm (a cap that covers the cervix) in combination with spermicide | 12 (4.1) |
| **The copper IUD (intrauterine device)** | **202 (69.2)** |
| The external condom | 78 (26.7) |
| Withdrawal before ejaculation / pull out method | 0 (0.0) |
| 3. What exactly does the morning-after pill do? | |
| It terminates a pregnancy | 25 (8.6) |
| It kills the sperm cells | 47 (16.1) |
| **It delays ovulation** | **155 (53.0)** |
| It ensures that menstruation comes | 65 (22.3) |
| 4. Which contraceptive method also protects against STDs? | |
| The contraceptive injection | 0 (0.0) |
| The contraceptive implant | 0 (0.0) |
| The copper IUD | 2 (0.7) |
| **The internal condom** | **290 (99.3)** |
| 5. How long can sperm stay alive in the vagina, uterus, and/or fallopian tube? | |
| 1–3 hours | 6 (2.1) |
| 24 hours | 29 (9.9) |
| **3–5 days** | **244 (83.5)** |
| 7–10 days | 13 (4.5) |
| 6. When during the menstrual cycle is a person most likely to become pregnant? | |
| During the period | 0 (0.0) |
| 3 days after the period is over | 20 (6.8) |
| **Two weeks before the period starts** | **245 (84.0)** |
| 3 days before the period starts | 27 (9.2) |
| 7. In which part of the body does a doctor or midwife place an IUD? | |
| Fallopian tube | 3 (1.0) |
| **Uterus** | **166 (56.9)** |
| Cervix | 123 (41.1) |
| Vagina | 0 (0.0) |
| Question 8. How long after stopping using oral contraception can someone get pregnant? | |

| | |
|---|---|
| **Immediately** | **265 (90.7)** |
| 1 month | 25 (8.6) |
| 3 months | 2 (0.7) |
| 6 months | 0 (0.0) |
